# Timing of Radiotherapy in Uveal Melanoma: A SEER-Based Comparison of Preoperative, Postoperative, and Intraoperative Approaches

**DOI:** 10.64898/2026.07.30.26359348

**Authors:** Mirsaeed Abdollahi, Fatemeh Razmjooei, Hamidreza Ashayeri, Farbod Semnani, Ali Jafarizadeh, Sepehr Fekrazad, Ryan Sameen Meshkin, J Fernando Arevalo

## Abstract

**Purpose:** To analyze the effect of radiotherapy timing relative to surgery on patients’ survival and mortality rates in uveal melanoma (UM).

**Design:** Retrospective cohort study using registry data from the Surveillance, Epidemiology, and End Results (SEER) database.

**Subjects:** A total of 1,198 patients with UM were extracted from the SEER database and divided into three groups: preoperative radiotherapy, intraoperative radiotherapy (IORT), and postoperative radiotherapy.

**Methods:** UM cases were extracted from the SEER database. Survival curves were generated based on the Kaplan-Meier method. An extended Cox regression was used to estimate the cancer-specific and all-cause mortality.

**Main Outcome Measures:** Overall survival (including median survival) and hazard reductions for cancer-specific and all-cause mortality.

**Results:** Preoperative radiotherapy revealed the highest median survival (4.93 years), followed by postoperative radiotherapy (4.76 years), and IORT (4.02 years). The preoperative strategy reduced the hazard of all-cause mortality by 42%, 50%, and 60% at 5-, 7-, and 10-year follow-up, respectively, compared with IORT. Findings were consistent in the cancer-specific sensitivity analysis (45%, 53%, and 62% reductions at the same time points). However, postoperative radiotherapy did not significantly reduce the cancer-specific mortality in comparison to IORT.

**Conclusions:** Preoperative radiotherapy was associated with higher median overall survival and lower all-cause and cancer-specific mortality than IORT over 10 years of follow-up. Postoperative radiotherapy showed intermediate outcomes that did not differ significantly from IORT. Prospective studies with more detailed data on the temporal relation of radiotherapy timing and surgery are needed to confirm these findings.

## 1. Introduction

Uveal melanoma (UM) is one of the deadliest primary intraocular malignancies originating from the melanocytes in the uveal tract. The age-adjusted incidence of UM is reported to be 5.1 per million per year ^1^. The median age for diagnosis of UM is 63 years, with a median overall survival (OS) of 154 months ^2^. UM is mainly seen in Caucasian populations, with no differences between males and females ^3,4^. The occurrence of UM is usually sporadic and rarely familial. UM differs from cutaneous melanoma in several aspects, the most important of which is the different clinical course and tumor genetics ^5,6^.

GNA11 and GNAQ are the most common mutations in UM and are thought to contribute to its pathogenesis in the early stages of disease ^7,8^. Other mutations, such as BAP1, SF3B1, and EIF1AX, are also observed with lower frequencies. Among these mutations, inactivation of BAP1 showed the highest metastatic potential ^9^. Other reported chromosomal abnormalities with increased risk of metastasis are 3p monosomy, 6p loss, 6q loss, 8p loss, and 8q gain ^10^. The UM’s location is also an essential factor in determining prognosis. The UM arising from the iris is detected 10-20 years earlier, which is associated with improved OS, whereas ciliary body and juxtapupillary tumors have the lowest survival rates. Other prognostic factors include a large basal diameter of the tumor > 16mm, thickness > 3mm, epithelioid cell type, and infiltrative lymphocytes in histopathology ^11^.

Current treatment options for UM are surgery, radiotherapy, and laser therapies ^11^. The most common radiotherapy techniques used for UM treatment include plaque brachytherapy, proton beam radiotherapy (PBR), and stereotactic radiosurgery ^12^. In a cohort study of 6,871 patients, radiotherapy alone was superior to surgery alone in improving long-term OS ^13^. A combination of surgical approaches with radiotherapy techniques has been proposed as a safe method to increase the OS ^14,15^. However, the data lack information on the optimal combination of these two approaches. In this retrospective study, we aim to compare the effects of different combinations of radiotherapy and surgery on patients’ survival.

## 2. Materials and Methods

### 2.1 Study design and participants

This retrospective cohort analysis, conducted using registry data, used the Surveillance, Epidemiology, and End Results (SEER) database accessed via SEER*Stat version 9.0.42.2. The SEER program provides comprehensive cancer incidence and survival data from population-based registries. Estimates based on the November 2024 submission indicate that the complete set of SEER 21 registries (excl IL) now covers approximately 46% of the U.S. population ^16^. We identified cases of uveal tract melanoma by selecting primary site codes C69.3 (choroid) and C69.4 (ciliary body), along with ICD-O-3 histology/behavior codes ranging from 8720/2 to 8790/3, in accordance with the International Classification of Diseases for Oncology. Iris melanomas (also coded as C69.4) were further identified using the American Joint Committee on Cancer (AJCC) ID (2018+) variable. Given the limited efficacy of chemotherapy in UM and its lack of routine clinical recommendation ^17^, patients who received chemotherapy were excluded from this study. By using the “RX Summ--Surg/Rad Seq (2006+)” and “Reason no cancer-directed surgery (2006+)” variables, the analysis focused on three distinct groups: those receiving radiation before surgery (Code 2), those receiving radiation after surgery (Code 3), and those receiving intraoperative radiation therapy (IORT) (Code 5) (**Error! Reference source not found.**).

**Figure 1.**
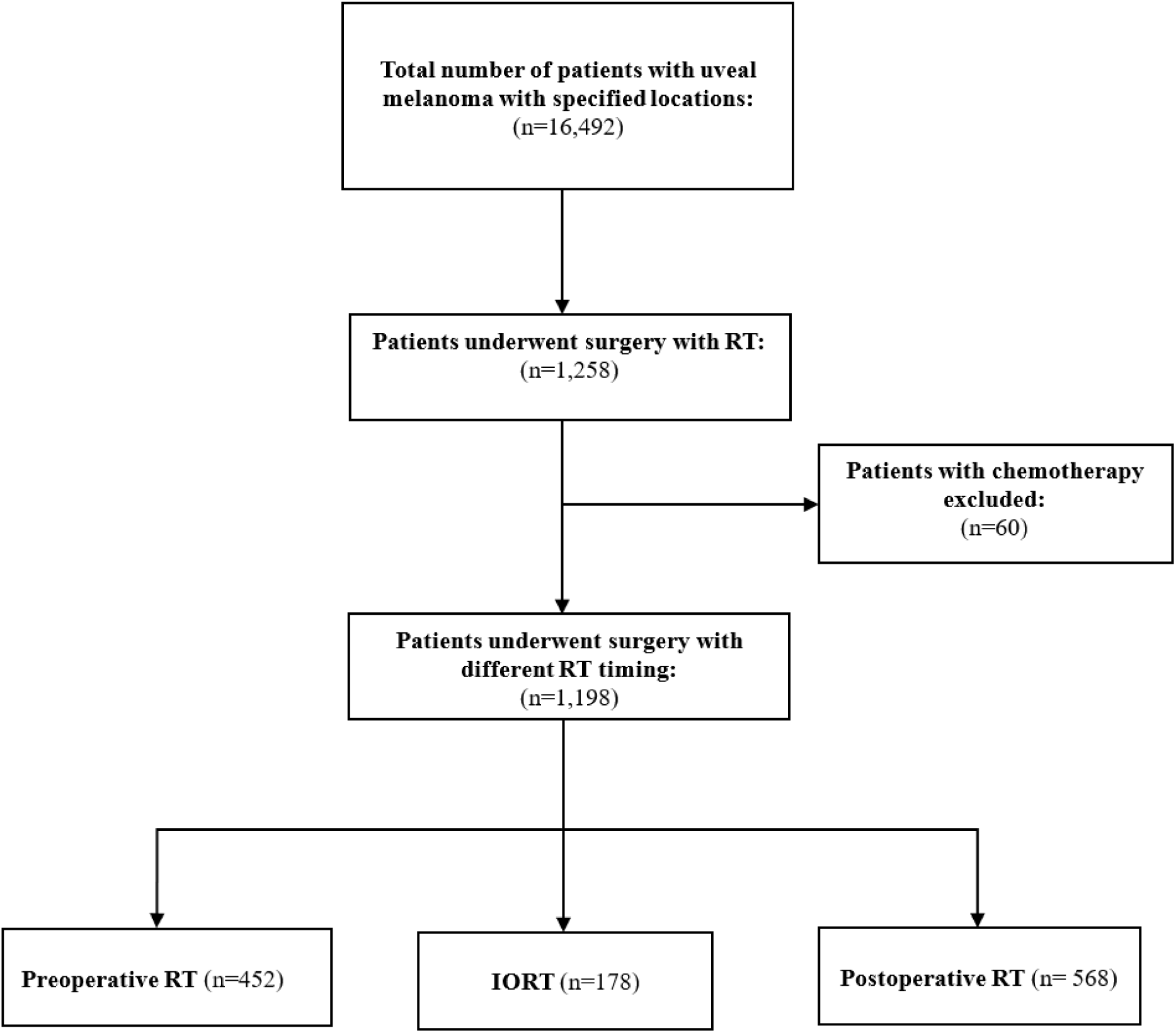
Flow diagram of study population inclusion

### 2.2 Variables

#### 2.2.1 Outcome variable

The primary outcome was OS at 3-, 5-, 7-, and 10-years following intervention for UM. OS was determined using follow-up duration and recorded death dates. Vital status was ascertained from the SEER vital status file, and patients were censored at the time of last known follow-up.

#### 2.2.2 Covariates & confounders

Demographic, treatment, and outcome variables were obtained from the SEER database. Demographic characteristics included age at diagnosis, sex, and race, with race recategorized as White, Black, and Asian. Clinical variables encompassed tumor location, laterality, histopathologic classification, tumor grade, disease stage, and the total number of in situ malignant tumors. Disease stage was defined according to the SEER Combined Summary Stage with Expanded Regional Codes.

According to existing literature, we considered some available variables—age, sex, race, primary tumor site, disease stage, and number of tumors—to adjust their potential effects when evaluating the comparative efficacy of the treatment groups ^18–21^. Due to the substantial proportion of missing or unspecified pathology and tumor grade information in the SEER dataset for UM, statistical adjustment for these variables was not feasible.

For minor confounders (generally defined as those with an odds ratio or hazard ratio <3 in relation to the outcome ^22^), statistical adjustment is considered sufficient to adequately control for their effects.

### 2.3 Statistical Analysis

Baseline characteristics were summarized using descriptive statistics. Continuous variables were reported as mean ± standard deviation (SD), and categorical variables were summarized as frequencies and percentages. OS was defined as the time from initial treatment to death from any cause. Survival analyses were performed using the Kaplan–Meier curves, and median survival times with 95% confidence intervals (CIs) were calculated for each treatment group. Survival probabilities at 3-, 5-, 7-, and 10-years were extracted for each radiotherapy timing group. At each time point, an overall Wald χ² test was performed to evaluate differences in survival probabilities among the three groups.

The proportional hazards (PH) assumption was assessed using Schoenfeld residuals and visually checked by plots, and was violated for the treatment group and primary tumor site (*p* < 0.05) (Figure S1). Therefore, an extended adjusted Cox model was fitted, with the primary tumor site handled by stratification to allow site-specific baseline hazards, and treatment effects modeled with time-varying coefficients (TVC). Alternative forms for time-varying treatment effects were compared using Akaike’s Information Criterion (AIC) ^23^. The Model incorporating linear time demonstrated a superior fit compared with the log-transformed model (12788.13 vs. 12793.72) and was therefore retained for final analyses. The primary effect size for survival analyses was the hazard ratio (HR). The log-hazard ratio for each treatment group relative to the reference group was modeled as a linear function of follow-up time. Specifically, the time-specific log hazard ratio until time *t* was defined as:

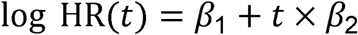

where *β*3_1_ represents the baseline (time-independent) treatment effect and *β*3_2_ denotes the coefficient of the interaction between treatment and follow-up time ^24^. Time-specific HRs and corresponding 95% confidence intervals (CI) were subsequently obtained by exponentiating the estimated log HRs. All statistical analyses were performed using STATA 18 (Stata Corp, College Station, TX).

Finally, as a sensitivity analysis, survival models were refitted using cancer-specific mortality (i.e., deaths attributed to UM) to evaluate the robustness of the primary findings based on all-cause mortality. Patients who died from causes other than UM were censored at the time of death. The same modeling strategy as in the primary analysis was applied, including Kaplan–Meier estimation and extended Cox regression with time-varying treatment effects, stratified by primary tumor site. Consistency between the main and sensitivity analyses was interpreted as evidence of robustness.

## 3. Results

### 3.1 Patient characteristics

A total of 1,198 patients were included in this study and were categorized by radiotherapy timing as IORT (n = 178), postoperative radiotherapy (n = 568), or preoperative radiotherapy (n = 452). Baseline demographic and clinicopathologic characteristics stratified by treatment group are summarized in Table 1. Age and sex distributions were similar across the three groups. However, the postoperative radiotherapy group included a higher proportion of ciliary body tumors (18.1% vs 6.7% in IORT and 5.8% in preoperative) and of regional or distant stage disease (16.9% vs 3.9% and 4.6%, respectively). These prognostically relevant imbalances were accounted for in the adjusted model through stratification by primary tumor site and inclusion of stage as a covariate. The mean age of patients and sex distribution were balanced across the three groups. The choroid was the predominant site of tumor origin, and most patients were diagnosed with localized disease. Most individuals had a single malignant tumor at the time of diagnosis.

**Table 1.** Baseline Characteristics of Patients with Uveal Melanoma by Treatment Group.

| Characteristic | IORT (n = 178) | Postoperative RT (n = 568) | Preoperative RT (n = 452) |
| --- | --- | --- | --- |
| Age, years (mean ± SD) | 62.2 ± 13.0 | 61.6 ± 14.0 | 60.7 ± 14.9 |
| Sex |  |  |  |
| Male | 94 (52.8%) | 288 (50.7%) | 226 (50.0%) |
| Female | 84 (47.2%) | 280 (49.3%) | 226 (50.0%) |
| Race |  |  |  |
| White | 174 (97.8%) | 540 (95.1%) | 446 (98.7%) |
| Asian | 0 (0.0%) | 14 (2.5%) | 2 (0.4%) |
| Black | 2 (1.1%) | 8 (1.4%) | 4 (0.9%) |
| Missing | 2 (1.1%) | 6 (1.1%) | 0 (0.0%) |
| Laterality |  |  |  |
| Left | 100 (56.2%) | 275 (48.4%) | 223 (49.3%) |
| Right | 78 (43.8%) | 292 (51.4%) | 228 (50.4%) |
| Paired site | 0 (0.0%) | 1 (0.2%) | 1 (0.2%) |
| Primary tumor site |  |  |  |
| Choroid | 165 (92.7%) | 461 (81.2%) | 425 (94.0%) |
| Ciliary body | 12 (6.7%) | 103 (18.1%) | 26 (5.8%) |
| **Iris** | 1 (0.6%) | 4 (0.7%) | 1 (0.2%) |
| **Tumor grade** |  |  |  |
| **Grade I–II** | 3 (1.7%) | 68 (12.0%) | 3 (0.7%) |
| **Grade III–IV** | 0 (0.0%) | 4 (0.7%) | 2 (0.4%) |
| **Unknown** | 175 (98.3%) | 496 (87.3%) | 447 (98.9%) |
| **Stage (Combined Summary Stage)** |  |  |  |
| **Localized** | 165 (92.7%) | 436 (76.8%) | 417 (92.3%) |
| **Regional involvement** | 7 (3.9%) | 84 (14.8%) | 16 (3.5%) |
| **Distant** | 0 (0.0%) | 12 (2.1%) | 5 (1.1%) |
| **Unknown** | 6 (3.4%) | 36 (6.3%) | 14 (3.1%) |
| **Histology** |  |  |  |
| **Melanoma, NOS** | 175 (98.3%) | 461 (81.2%) | 416 (92.0%) |
| **Spindle cell** | 2 (1.1%) | 53 (9.3%) | 14 (3.1%) |
| **Epithelioid** | 0 (0.0%) | 11 (1.9%) | 6 (1.3%) |
| **Mixed Epithelioid and Spindle** | 0 (0.0%) | 40 (7.0%) | 10 (2.2%) |
| **Amelanotic** | 1 (0.6%) | 3 (0.5%) | 5 (1.1%) |
| **Superficial Spreading** | 0 (0.0%) | 0 (0.0%) | 1 (0.2%) |
| **Number of malignant tumors** |  |  |  |
| **1** | 132 (74.2%) | 424 (74.7%) | 311 (68.8%) |
| **2** | 36 (20.2%) | 112 (19.7%) | 109 (24.1%) |
| **≥3** | 10 (5.6%) | 32 (5.6%) | 32 (7.1%) |

### 3.2 Survival analysis

Mean follow-up duration differed across treatment groups, with the shortest follow-up observed in the IORT group (4.77 ± 3.5 years), followed by the postoperative radiotherapy group (5.95 ± 4.5 years), and the longest follow-up in the preoperative radiotherapy group (6.68 ± 5.2 years). As shown in Kaplan-Meier, survival curves were significantly different across radiotherapy-timing groups using the Wilcoxon–Breslow–Gehan test (p=0.001) (**Error! Reference source not found.**).

**Figure 2.**
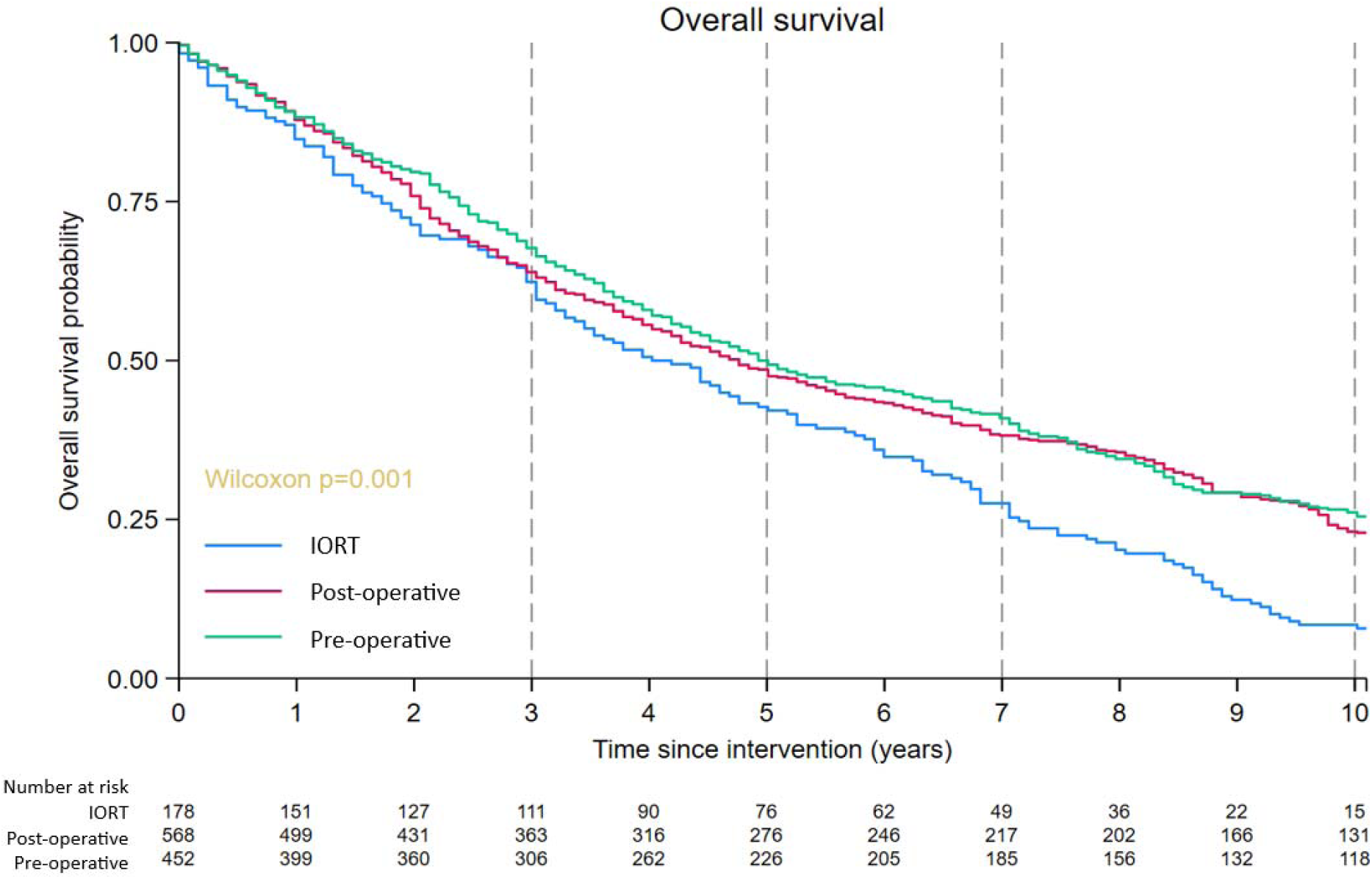
Kaplan–Meier curves comparing survival in three groups: preoperative radiotherapy, postoperative radiotherapy, and intraoperative radiation therapy (IORT)

Table 2 summarizes OS estimates at prespecified time points by radiotherapy timing. At year 3, OS was 62.4% (95% CI, 54.8–69.0) in the IORT group, 63.9% (95% CI, 59.8–67.7) in the postoperative radiotherapy group, and 67.7% (95% CI, 63.2–71.8) in the preoperative radiotherapy group, with no statistically significant difference among groups (p = 0.315). Similarly, 5-year survival did not differ significantly across treatment strategies (IORT: 42.7%; postoperative radiotherapy: 48.6%; preoperative radiotherapy: 50.0%; p = 0.244). In contrast, statistically significant differences emerged at later time points. At year 7, survival was lowest in the IORT group (27.5%; 95% CI, 21.2–34.2) compared with the postoperative (38.2%; 95% CI, 34.2–42.2) and preoperative radiotherapy groups (40.9%; 95% CI, 36.4–45.4), with a significant overall difference (p = 0.004). This divergence was more pronounced at year 10, where survival estimates were 8.4% (95% CI, 4.9–13.1) for IORT, 23.1% (95% CI, 19.7–26.6) for postoperative radiotherapy, and 26.1% (95% CI, 22.1–30.2) for preoperative radiotherapy (p < 0.001). Median OS was shortest in the IORT group (4.02 years; 95% CI, 3.3–4.9), intermediate in the postoperative radiotherapy group (4.76 years; 95% CI, 4.2–5.4), and longest in the preoperative radiotherapy group (4.93 years; 95% CI, 4.4–6.0).

**Table 2.**
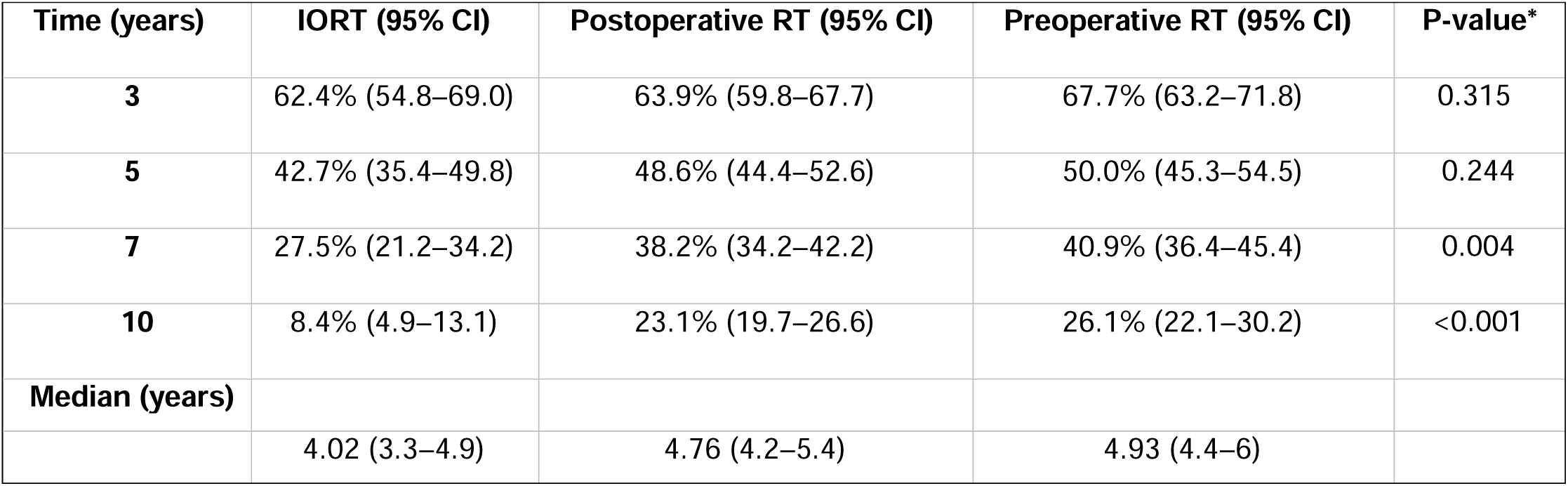
Survival rate at specific time points and median survival time.

| Time (years) | IORT (95% CI) | Postoperative RT (95% CI) | Preoperative RT (95% CI) | P-value* |
| --- | --- | --- | --- | --- |
| 3 | 62.4% (54.8–69.0) | 63.9% (59.8–67.7) | 67.7% (63.2–71.8) | 0.315 |
| 5 | 42.7% (35.4–49.8) | 48.6% (44.4–52.6) | 50.0% (45.3–54.5) | 0.244 |
| 7 | 27.5% (21.2–34.2) | 38.2% (34.2–42.2) | 40.9% (36.4–45.4) | 0.004 |
| 10 | 8.4% (4.9–13.1) | 23.1% (19.7–26.6) | 26.1% (22.1–30.2) | <0.001 |
| Median (years) |  |  |  |  |
|  | 4.02 (3.3–4.9) | 4.76 (4.2–5.4) | 4.93 (4.4–6) |  |

*\*P-values were derived from pointwise Wald $\chi^2$ tests*

In the stratified extended adjusted Cox model, allowing treatment effects to vary linearly with time, the OS of different radiotherapy methods was time-dependent (Table 3). After 5 years, preoperative radiotherapy was significantly superior to IORT with a lower hazard of death (HR = 0.58; 95% CI, 0.34–0.97; p = 0.042). Similarly, after 7 years, preoperative radiotherapy continued to demonstrate a significant survival advantage compared with IORT (HR = 0.50; 95% CI, 0.27–0.92; p = 0.027) and remained evident after 10 years, where preoperative radiotherapy was significantly superior in hazard (HR = 0.40; 95% CI, 0.19–0.84; p = 0.016). However, during the entire period, the superiority of postoperative radiotherapy over IORT remained non-significant.

**Table 3.** Time-specific adjusted hazard ratios for all-cause mortality from the stratified extended Cox mod.

| Time (years) | IORT | Postoperative RT HR (95% CI) | p-value | Preoperative RT HR (95% CI) | p-value |
| --- | --- | --- | --- | --- | --- |
| 3 | Ref. | 0.80 (0.52–1.22) | 0.305 | 0.67 (0.43–1.03) | 0.072 |
| 5 | Ref. | 0.73 (0.43–1.23) | 0.240 | 0.58 (0.34–0.97) | 0.042 |
| 7 | Ref. | 0.67 (0.36–1.24) | 0.204 | 0.50 (0.27–0.92) | 0.027 |
| 10 | Ref. | 0.59 (0.28–1.25) | 0.167 | 0.40 (0.19–0.84) | 0.016 |

### 3.3 Sensitivity analysis

According to Table 4, sensitivity analyses using cancer-specific mortality yielded results consistent with the primary analysis (**Error! Reference source not found.**).

**Figure 3.**
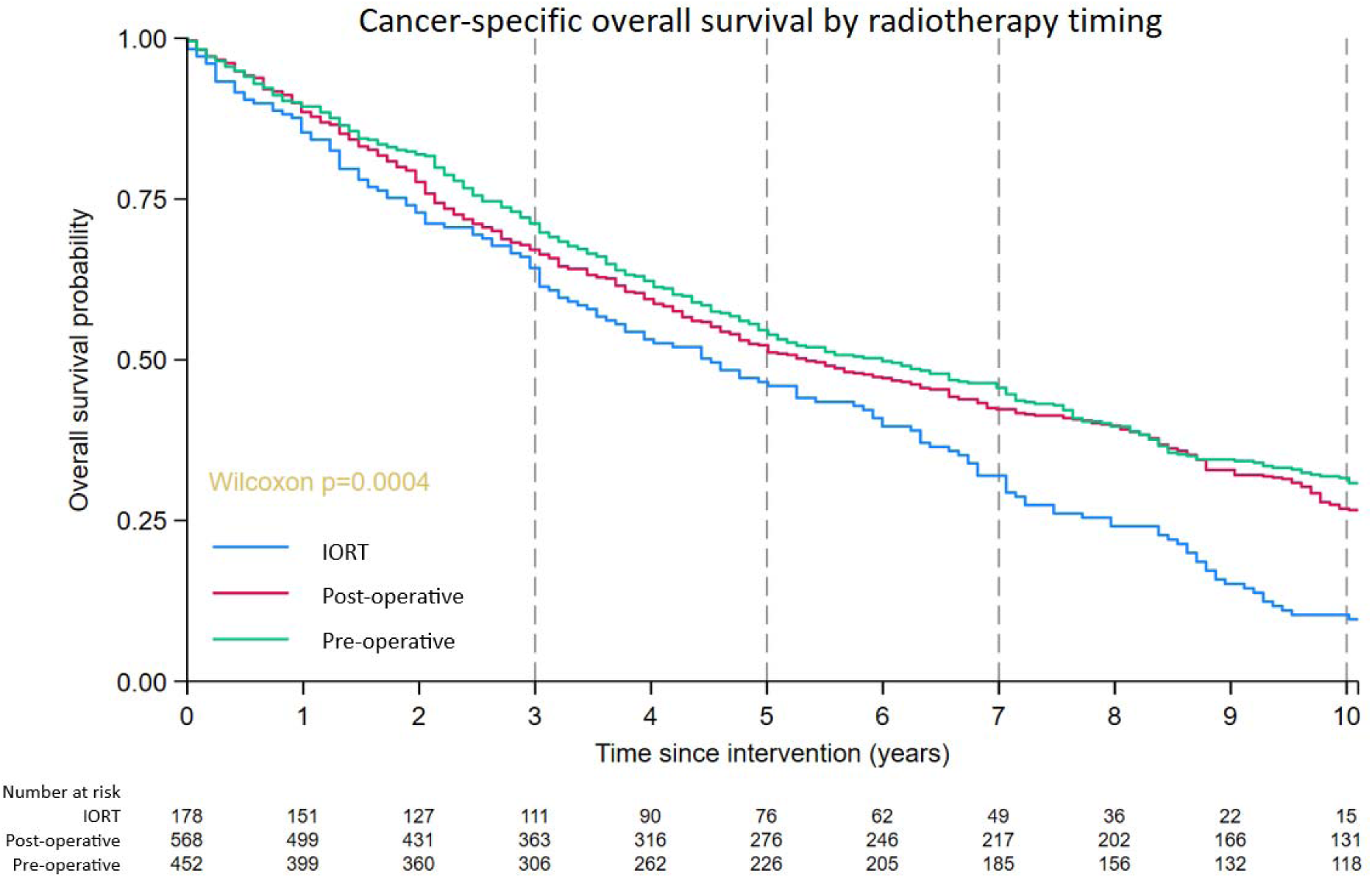
Kaplan–Meier curves comparing cancer-specific survival in three groups; preoperative radiotherapy, postoperative radiotherapy, and intraoperative radiation therapy (IORT)

**Table 4.** Time-specific adjusted hazard ratios for cancer-specific mortality (sensitivity analysis)

| Time (years) | IORT | postoperative RT HR (95% CI) | preoperative RT HR (95% CI) |
| --- | --- | --- | --- |
| 3 | Ref. | 0.80 (0.51–1.24) | 0.65 (0.41–1.01) |
| 5 | Ref. | 0.73 (0.42–1.25) | 0.55 (0.32–0.96) |
| 7 | Ref. | 0.67 (0.35–1.27) | 0.47 (0.25–0.91) |
| 10 | Ref. | 0.59 (0.27–1.30) | 0.38 (0.17–0.84) |

## 4. Discussion

Our study indicated that preoperative radiotherapy was associated with higher OS. Based on the adjusted hazard ratios, preoperative radiotherapy reduced the hazard of all-cause mortality by 42%, 50%, and 60% compared with IORT at 5-, 7-, and 10-year follow-up, respectively; the cancer-specific sensitivity analysis showed comparable reductions (45%, 53%, and 62%). Preoperative radiotherapy was superior to IORT for the treatment of localized choroidal UM, which comprised most of our sample.

However, postoperative results should be interpreted with extreme caution. Although postoperative radiotherapy did not reduce overall or cancer-specific mortality compared with IORT during the entire follow-up period (p-value >0.05), its Kaplan–Meier curve crossed multiple times with preoperative radiotherapy. These crossings and OS overlap in Table 2, suggest possibility of a non-significant difference between preoperative and postoperative radiotherapy. These mixing results may reflect from the lack of an exact temporal relationship between radiotherapy and surgery in the preoperative and postoperative groups. Another important factor to consider is the relatively lower OS among all three groups compared with the 154 months reported by Tuz Zahra et al. ^2^. This may indicate that patients in our cohort had a more aggressive or high-grade UM, requiring both surgical and RT interventions.

Preoperative therapy does not lead to a significant reduction in mortality rates in the three years; however, it may have benefits thereafter. This is maybe due to the timing of tumor recurrence. Boker et al. ^25^, found that neoadjuvant PBR with transscleral resection reduced tumor recurrence but did not reduce the need for enucleation. Similarly, Willerding et al. ^26^ reported a reduction in local recurrence. Tumor regression following the radiotherapy ^27^ may also enable the surgeon to place the margin closer and reduce the size of the surgical coloboma ^26^.

Although tumors can recur locally as early as 1 month, the rate in the first 3 years after treatment has been reported to be 3-3.8% ^28,29^. Siebel et al. ^29^ reported a median time to recurrence of 17.5 months in patients with UM, and Caujolle et al. ^30^ reported that most recurrences happened in the first 3 years. However, neither study has reported whether patients received surgical intervention in addition to PBR, which may be associated with earlier local recurrence ^29,30^. A study of 4,196 patients treated with radiotherapy reports a median time to local recurrence of 30.5 months ^31^, which is longer than previously reported ^29^. In a study by Kolandajan et al. ^32^ median time to UM systemic recurrence was 3.25 years, exceeding the 3-year follow-up period. However, this study was conducted in patients with metastatic UM, whereas our study mostly included localized UM ^32^.

In a retrospective analysis of data from 6871 patients with choroidal UM, Wu et al. found that radiotherapy alone was superior to surgery or to surgery combined with radiotherapy ^13^. They report that the combination therapy had superior OS among patients under 25 years old, with small tumors, or stages as N1. The addition of radiotherapy to surgery had a similar OS in patients with T4 choroidal UM to surgery alone. The combination therapy also showed similar OS with the radiation-only group in T1, T3, M1, and large tumors. They attribute their findings to the possibility of tumor dissemination into the bloodstream during surgical manipulation ^33^. In their study, only 8.3% received a combination of surgery and radiotherapy, and 71.4% received radiotherapy only ^13^. They also didn’t report the timing of radiotherapy in relation to surgery ^13^.

Radiotherapy exerts anti-tumor effects by inducing DNA damage, disrupting cell membranes, and releasing lysosomal enzymes ^34^. Brachytherapy is performed using either gamma-emitting iodine-125 or beta-emitting ruthenium-106 plaques ^35^. It is suggested by the Collaborative Ocular Melanoma Study (COMS) and commonly used at a dose of 85 Gy to the tumor apex for medium-sized tumors ^36^; however, some studies suggest that lower doses may be sufficient ^37^. PBR uses pencil beam scanning to deliver protons to the target tumor at a dose of 56-60 Gy, divided into 4 sessions and 50-70 Gy, divided into 5 sessions ^38^. Because of the Bragg peak effect, PBR causes less damage to the surrounding tissue compared to brachytherapy ^12^. In stereotactic radiotherapy, photons are focused on the target tissue from different directions. The Gamma Knife and the linear particle accelerator (LINAC)-based devices are used in this method ^39^.

Ocular radiotherapy may have some side effects, such as scleritis, scleral necrosis, radiation maculopathy, optic neuropathy, vitreous hemorrhage, retinal detachment, secondary glaucoma, neovascular glaucoma, and toxic tumor syndrome ^40^. The prevalence of each complication is associated with the type of radiotherapy used and the tumor location ^40,41^. For example, the anteriorly located UM increases the risk of glaucoma ^41^, while UM near the optic disk increases the risk of optic neuropathy and vitreous hemorrhage ^40,42^. Toxic tumor syndrome is one of the late and severe complications of radiotherapy due to secondary vasculopathy ^43^. Diagnosis of this condition is crucial as it threatens vision, and patients may require additional surgery, such as vitreoretinal surgery or enucleation ^43,44^. There are also newer treatment options for UM, including transpupillary thermotherapy ^45^, immune checkpoint inhibitors, mitogen-activated protein kinase inhibitors, and tyrosine kinase inhibitors ^46–48^.

## 5. Limitations

This study has several limitations. First, although plaque brachytherapy is the most common radiotherapy modality used with surgery, detailed data on radiation technique were not consistently recorded in the SEER database, particularly before 2018. As a result, we were unable to determine the contribution of differences in radiotherapy modalities to the survival differences observed across preoperative, postoperative, and intraoperative groups. In addition, because plaque brachytherapy is delivered during a surgical episode, some plaque-treated cases may have been classified as intraoperative rather than as sequential surgery and radiation, so the timing groups may partly reflect coding practice rather than true clinical sequencing. Second, the distance of choroidal UM to the macula and optic nerve was not included. This distance is important because a lower dose near the respective locations may be used, affecting the OS. Third, histology data were missing for most tumors in our study. Fourth, the lack of surgery details and approaches in the SEER database. These crucial details should be considered and used to stratify the analysis and identify the optimal treatment strategy for each condition. Fifth, the exact interval between radiotherapy and surgery is not recorded; SEER captures only the sequence (before, during, or after surgery). Because follow-up was anchored at the initial treatment and group membership required receipt of both treatments, patients in the preoperative and postoperative groups had to survive from the first to the second treatment, introducing a period of immortal time relative to the intraoperative group. The interval between radiotherapy and surgery in combined treatment of uveal melanoma is typically short, so the resulting bias is expected to be modest, but it cannot be quantified with the available data and may contribute to the apparently poorer survival of the IORT group. Another limitation is the lack of data on radiotherapy-related side effects during the follow-up period. Lastly, our study had a retrospective design. For future studies, we recommend including more detailed data on histology, location, and surgery, and employing a prospective design to better inform treatment plans for UM.

## 6. Conclusion

In conclusion, the choice of treatment plan for UM requires careful consideration of patient- and tumor-specific characteristics. The side effects of treatment, the patient’s quality of life, and vision are also important to consider. This study supports consideration of preoperative radiotherapy over IORT, particularly in localized choroidal UM; postoperative outcomes were intermediate and did not differ significantly from IORT. Prospective studies with more details about the temporal relation of RT timing and surgery are needed to confirm whether these associations reflect a causal effect of treatment timing.

## Data Availability

All data produced are available online at https://seer.cancer.gov.

## Acknowledgement

None.

## CRediT authorship contribution statement

**Mirsaeed Abdollahi:** Conceptualization, Methodology, Formal analysis, Investigation, Writing – Original Draft, Writing – Review & Editing. **Fatemeh Razmjooei:** Visualization, Writing – Original Draft, Writing – Review & Editing. **Hamidreza Ashayeri:** Writing – Original Draft, Writing – Review & Editing. **Farbod Semnani**: Methodology, Writing – Review & Editing. **Ali Jafarizadeh:** Project administration, Supervision, Formal analysis, Writing – Original Draft, Writing – Review & Editing. **Sepehr Fekrazad:** Validation, Writing – Review & Editing. **Ryan Sameen Meshkin:** Validation, Writing – Review & Editing. **J. Fernando Arevalo:** Project administration, Supervision, Writing – Review & Editing. Mirsaeed Abdollahi and Fatemeh Razmjooei contributed equally to this work and share first authorship. All authors read and approved the final manuscript.

## Appendices

**Figure S1.**
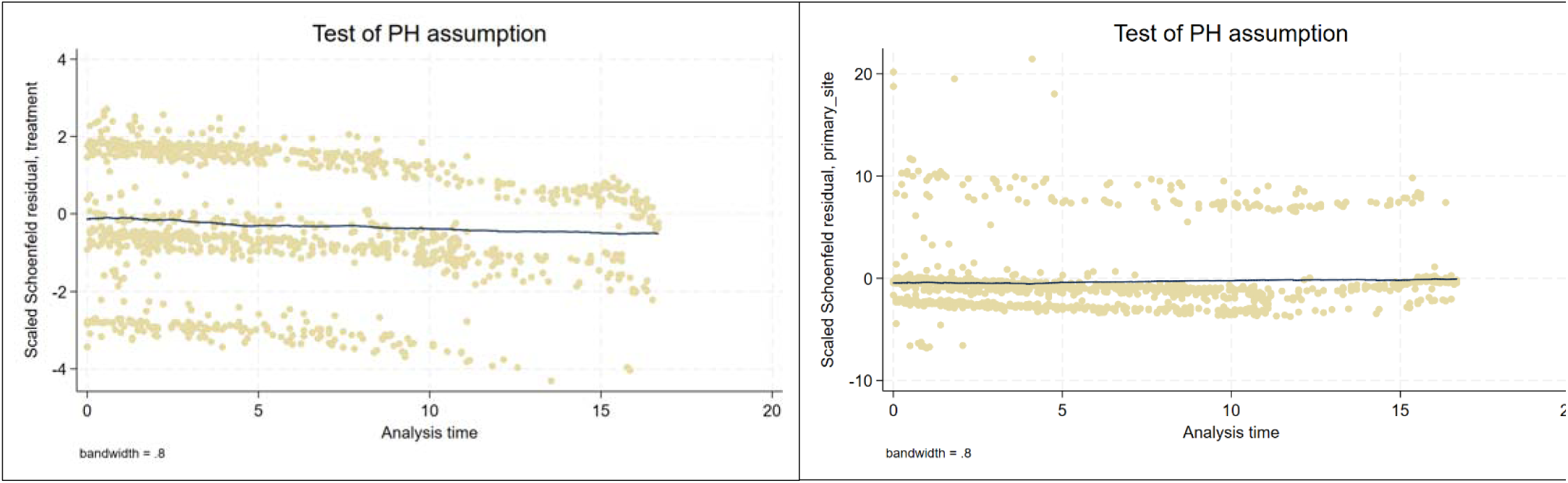
Schoenfeld residuals plot for two variables violating the PH assumption

## References

1. Kaliki S, Shields CL. Uveal melanoma: relatively rare but deadly cancer. Eye. 2017;31(2):241–257.

2. Tuz Zahra F, Ali H, Mohsin F, Bajwa S, Farhan SH, Jaglal MV. Trends in mortality, incidence, and survival of uveal melanoma: A SEER-based retrospective analysis. Journal of Clinical Oncology. 2025/06/01 2025;43(16_suppl):e21581–e21581. doi:10.1200/JCO.2025.43.16_suppl.e21581

3. Sener H, Bansal R, Shields JA, Shields CL. Influence of race on uveal melanoma metastasis-free survival: a matched comparative analysis between African American and Caucasian American cohorts. Eye. 2025:1–3.

4. Damato B, Coupland S. Differences in uveal melanomas between men and women from the British Isles. Eye. 2012;26(2):292–299.

5. Helgadottir H, Höiom V. The genetics of uveal melanoma: current insights. The application of clinical genetics. 2016:147–155.

6. van der Kooij MK, Speetjens FM, van der Burg SH, Kapiteijn E. Uveal versus cutaneous melanoma; same origin, very distinct tumor types. Cancers. 2019;11(6):845.

7. Onken MD, Worley LA, Long MD, et al. Oncogenic mutations in GNAQ occur early in uveal melanoma. Investigative ophthalmology & visual science. 2008;49(12):5230–5234.

8. Van Raamsdonk CD, Griewank KG, Crosby MB, et al. Mutations in GNA11 in uveal melanoma. New England Journal of Medicine. 2010;363(23):2191–2199.

9. Decatur CL, Ong E, Garg N, et al. Driver mutations in uveal melanoma: associations with gene expression profile and patient outcomes. JAMA ophthalmology. 2016;134(7):728–733.

10. Vaquero-Garcia J, Lalonde E, Ewens KG, et al. PRiMeUM: a model for predicting risk of metastasis in uveal melanoma. Investigative ophthalmology & visual science. 2017;58(10):4096–4105.

11. Kulbay M, Marcotte E, Remtulla R, et al. Uveal melanoma: Comprehensive review of its pathophysiology, diagnosis, treatment, and future perspectives. Biomedicines. 2024;12(8):1758.

12. Bai H, Bosch JJ, Heindl LM. Current management of uveal melanoma: A review. Clinical & experimental ophthalmology. 2023;51(5):484–494.

13. Wu Y, Shi L, Ye Z, Zhou Y, Wang F, Zhang Y. Radiotherapy has a survival advantage over surgery in patients with choroidal melanoma: a retrospective cohort study of 6,871 patients. Front Surg. 2025;12:1577775. doi:10.3389/fsurg.2025.1577775

14. Relimpio-López I, Garrido-Hermosilla AM, Espejo F, et al. Clinical outcomes after surgical resection combined with brachytherapy for uveal melanomas. Journal of Clinical Medicine. 2022;11(6):1616.

15. Suesskind D, Scheiderbauer J, Buchgeister M, et al. Retrospective evaluation of patients with uveal melanoma treated by stereotactic radiosurgery with and without tumor resection. JAMA ophthalmology. 2013;131(5):630–637.

16. Surveillance E, and End Results (SEER) Program. Data from: SEER*Stat Database: Incidence – SEER Research Limited-Field Data, 21 Registries (excluding Illinois), Nov 2024 Submission (2000–2022), Linked to County Attributes – Time Dependent (1990–2023) Income/Rurality, 1969–2023 Counties. 2025. Bethesda, MD. Deposited Released April 2025; based on the November 2024 submission.

17. Scoles S, Ganesh S, Yamada KH. Current Therapies and Potential Strategies for Uveal Melanoma. Drugs and Drug Candidates. 2025;4(2):14.

18. Rao YJ, Sein J, Badiyan S, et al. Patterns of care and survival outcomes after treatment for uveal melanoma in the post-coms era (2004-2013): a surveillance, epidemiology, and end results analysis. Journal of contemporary brachytherapy. 2017;9(5):453–465.

19. Luo J, Zhang C, Yang Y, et al. Characteristics, treatments, and survival of Uveal Melanoma: a comparison between Chinese and American cohorts. Cancers. 2022;14(16):3960.

20. Mellen PL, Morton SJ, Shields CL. American joint committee on cancer staging of uveal melanoma. Oman J Ophthalmol. May 2013;6(2):116–8. doi:10.4103/0974-620x.116652

21. Dogrusöz M, Jager MJ, Damato B. Uveal melanoma treatment and prognostication. Asia-Pacific Journal of Ophthalmology. 2017;6(2):186–196.

22. Taubes G. Epidemiology faces its limits: the search for subtle links between diet, lifestyle, or environmental factors and disease is an unending source of fear—but often yields little certainty. Science. 1995;269(5221):164–169.

23. Burnham KP, Anderson DR. Model selection and multimodel inference: a practical information-theoretic approach. Springer; 2002.

24. Therneau TM, Grambsch PM. The cox model. Modeling survival data: extending the Cox model. Springer; 2000:39–77.

25. Böker A, Pilger D, Cordini D, et al. Neoadjuvant proton beam irradiation vs. adjuvant ruthenium brachytherapy in transscleral resection of uveal melanoma. Graefe’s Archive for Clinical and Experimental Ophthalmology. 2018;256(9):1767–1775.

26. Willerding GD, Cordini D, Moser L, Krause L, Foerster MH, Bechrakis NE. Neoadjuvant proton beam irradiation followed by transscleral resection of uveal melanoma in 106 cases. Br J Ophthalmol. Apr 2016;100(4):463–7. doi:10.1136/bjophthalmol-2015-307095

27. Fang R, Wang H, Li Y, Liu Y-M, Wei W-B. Regression patterns of uveal melanoma after iodine-125 plaque brachytherapy. BMC Ophthalmology. 2021/03/16 2021;21(1):137. doi:10.1186/s12886-021-01898-3

28. The Ophthalmic Oncology Task F, Gallie BL, Simpson ER, et al. Local Recurrence Significantly Increases the Risk of Metastatic Uveal Melanoma. Ophthalmology. 2016;123(1):86–91. doi:10.1016/j.ophtha.2015.09.014

29. Seibel I, Cordini D, Rehak M, et al. Local Recurrence After Primary Proton Beam Therapy in Uveal Melanoma: Risk Factors, Retreatment Approaches, and Outcome. American Journal of Ophthalmology. 2015;160(4):628–636. doi:10.1016/j.ajo.2015.06.017

30. Caujolle J-P, Paoli V, Chamorey E, et al. Local Recurrence After Uveal Melanoma Proton Beam Therapy: Recurrence Types and Prognostic Consequences. International Journal of Radiation Oncology*Biology*Physics. 2013/04/01/ 2013;85(5):1218–1224. 10.1016/j.ijrobp.2012.10.005

31. Lane AM, Hartley C, Go AK, Wu F, Gragoudas ES, Kim IK. Survival of patients with recurrent uveal melanoma after treatment with radiation therapy. Br J Ophthalmol. May 21 2024;108(5):729–734. doi:10.1136/bjo-2022-323133

32. Kolandjian NA, Wei C, Patel SP, et al. Delayed systemic recurrence of uveal melanoma. Am J Clin Oncol. Oct 2013;36(5):443–9. doi:10.1097/COC.0b013e3182546a6b

33. Beasley AB, Chen FK, Isaacs TW, Gray ES. Future perspectives of uveal melanoma blood based biomarkers. Br J Cancer. Jun 2022;126(11):1511–1528. doi:10.1038/s41416-022-01723-8

34. Groenewald C, Konstantinidis L, Damato B. Effects of radiotherapy on uveal melanomas and adjacent tissues. Eye. 2013/02/01 2013;27(2):163–171. doi:10.1038/eye.2012.249

35. Takiar V, Voong KR, Gombos DS, et al. A choice of radionuclide: Comparative outcomes and toxicity of ruthenium-106 and iodine-125 in the definitive treatment of uveal melanoma. Practical Radiation Oncology. 2015/05/01/ 2015;5(3):e169–e176. 10.1016/j.prro.2014.09.005

36. The COMS randomized trial of iodine 125 brachytherapy for choroidal melanoma: V. Twelve-year mortality rates and prognostic factors: COMS report No. 28. Arch Ophthalmol. Dec 2006;124(12):1684–93. doi:10.1001/archopht.124.12.1684

37. Echegaray JJ, Bechrakis NE, Singh N, Bellerive C, Singh AD. Iodine-125 Brachytherapy for Uveal Melanoma: A Systematic Review of Radiation Dose. Ocul Oncol Pathol. Sep 2017;3(3):193–198. doi:10.1159/000455872

38. Chan AW, Lin H, Yacoub I, Chhabra AM, Choi JI, Simone CB, 2nd. Proton Therapy in Uveal Melanoma. Cancers (Basel). Oct 16 2024;16(20)doi:10.3390/cancers16203497

39. Semeniuk O, Yu E, Rivard MJ. Current and Emerging Radiotherapy Options for Uveal Melanoma. Cancers. 2024;16(5):1074.

40. Banou L, Tsani Z, Arvanitogiannis K, Pavlaki M, Dastiridou A, Androudi S. Radiotherapy in Uveal Melanoma: A Review of Ocular Complications. Curr Oncol. Jul 3 2023;30(7):6374–6396. doi:10.3390/curroncol30070470

41. Kim EA, Salazar D, McCannel CA, et al. Glaucoma After Iodine-125 Brachytherapy for Uveal Melanoma: Incidence and Risk Factors. J Glaucoma. Jan 2020;29(1):1–10. doi:10.1097/ijg.0000000000001393

42. Riechardt AI, Cordini D, Willerding GD, et al. Proton beam therapy of parapapillary choroidal melanoma. Am J Ophthalmol. Jun 2014;157(6):1258–65. doi:10.1016/j.ajo.2014.02.032

43. Romano MR, Catania F, Confalonieri F, et al. Vitreoretinal Surgery in the Prevention and Treatment of Toxic Tumour Syndrome in Uveal Melanoma: A Systematic Review. Int J Mol Sci. Sep 17 2021;22(18)doi:10.3390/ijms221810066

44. Padley T, Hussain R, Eleuteri A, Chou HD, Groenewald C, Heimann H. Vitreoretinal Surgery for Intraocular Complications Following Radiotherapy Treatment of Uveal Melanoma. Cancers. 12/27 2025;18:95. doi:10.3390/cancers18010095

45. Desjardins L, Lumbroso-Le Rouic L, Levy-Gabriel C, et al. Combined proton beam radiotherapy and transpupillary thermotherapy for large uveal melanomas: a randomized study of 151 patients. Ophthalmic Res. 2006;38(5):255–60. doi:10.1159/000094834

46. Seth R, Agarwala SS, Messersmith H, et al. Systemic Therapy for Melanoma: ASCO Guideline Update. Journal of Clinical Oncology. 2023/10/20 2023;41(30):4794–4820. doi:10.1200/JCO.23.01136

47. Carvajal RD, Schwartz GK, Mann H, Smith I, Nathan PD. Study design and rationale for a randomised, placebo-controlled, double-blind study to assess the efficacy of selumetinib (AZD6244; ARRY-142886) in combination with dacarbazine in patients with metastatic uveal melanoma (SUMIT). BMC Cancer. Jun 10 2015;15:467. doi:10.1186/s12885-015-1470-z

48. Valsecchi ME, Orloff M, Sato R, et al. Adjuvant Sunitinib in High-Risk Patients with Uveal Melanoma: Comparison with Institutional Controls. Ophthalmology. Feb 2018;125(2):210–217. doi:10.1016/j.ophtha.2017.08.017

